# Machine learning model estimating number of COVID-19 infection cases over coming 24 days in every province of South Korea (XGBoost and MultiOutputRegressor)

**DOI:** 10.1101/2020.05.10.20097527

**Authors:** Yoshiro Suzuki, Ayaka Suzuki, Shun Nakamura, Toshiko Ishikawa, Akira Kinoshita

**Affiliations:** Tokyo Institute of Technology; Ernst & Young ShinNihon LLC

## Abstract

We built a machine learning model (ML model) which input the number of daily infection cases and the other information related to COVID-19 over the past 24 days in each of 17 provinces in South Korea, and output the total increase in the number of infection cases in each of 17 provinces over the coming 24 days. We employ a combination of XGBoost and MultiOutputRegressor as machine learning model (ML model). For each province, we conduct a binary classification whether our ML model can classify provinces where total infection cases over the coming 24 days is more than 100. The result is Sensitivity = 3/3 = 100%, Specificity = 11/14 = 78.6%, False Positive Rate = 3/11 = 21.4%, Accuracy = 14/17 = 82.4%. Sensitivity = 100% means that we did not overlook the three provinces where the number of COVID-19 infection cases increased by more than100. In addition, as for the provinces where the actual number of new COVID-19 infection cases is less than 100, the ratio (Specificity) that our ML model can correctly estimate was 78.6%, which is relatively high. From the above all, it is demonstrated that there is a sufficient possibility that our ML model can support the following four points. (1) Promotion of behavior modification of residents in dangerous areas, (2) Assistance for decision to resume economic activities in each province, (3) Assistance in determining infectious disease control measures in each province, (4) Search for factors that are highly correlated with the future increase in the number of COVID-19 infection cases.

## 1. Introduction

The new-type corona virus (SARS-CoV-2), which suddenly appeared at the end of 2019, caused global epidemic of COVID-19 in 2020. As of May 2020, industry, government, and academia from each country are cooperating to take measures to prevent the spreading of COVID-19 infection and develop therapeutic drugs.

This is the third time in this century that humans have been threatened by corona virus. The first was SARS in 2003, the second was MERS in 2012, and the third is COVID-19. Comparing SARS and MERS with COVID-19, it can be said that computer science including artificial intelligence (AI) and machine learning (ML) has made great progress. In fact, many computer science approaches have been developed to prevent the spreading of COVID-19 infection. For example, AI and ML are used for infection spreading analysis, drug discovery assistance, automatic diagnosis, diagnosis assistance, social trend analysis, and infection route analysis.

This paper proposes an ML technology for infection spreading analysis. To be more specific, we input the number of COVID-19 infection cases per day and other related information in each of 17 provinces in South Korea for the last 24 days, and output the total increase in the number of infection cases in each of 17 provinces over the coming 24 days. The result of conducting binary classification whether the total number of COVID-19 infection cases exceeds 100 over coming 24 days is Sensitivity = 3/3 = 100%, Specificity = 11/14 = 78.6%, False Positive Rate = 3 / 11 = 21.4%, Accuracy = 14/17 = 82.4%.

The existing ML for analyzing spreading of infectious diseases is introduced in Section 2. However, at present (May 10th, 2020), there is no ML that can estimate the number of COVID-19 infection cases in each province in South Korea with the above-mentioned high accuracy. From the above all, it is demonstrated that there is a sufficient possibility that our ML model can support the following four points. (1) Promotion of behavior modification of residents in dangerous areas, (2) Assistance for decision to resume economic activities in each region, (3) Assistance in determining infectious disease control measures in each region, (4) Search for factors that are highly correlated with the future increase in the number of COVID-19 infection cases.

## 2. Related works

In **Table 1**, we introduce the existing works about ML models which predict the spreading of COVID-19 infection. Also, we show the novelty points of our ML model compared to the existing works.

**Table 1.**
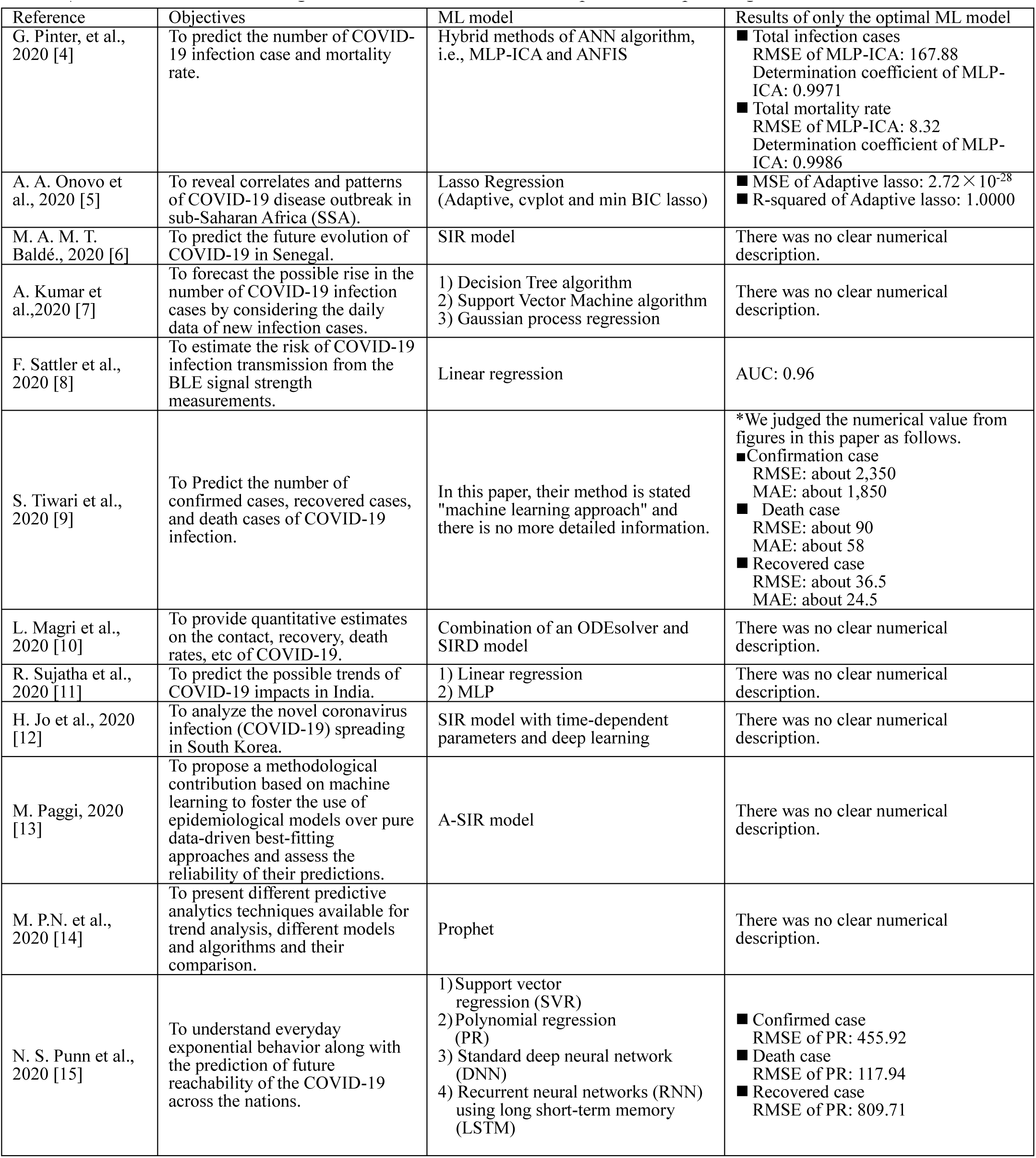
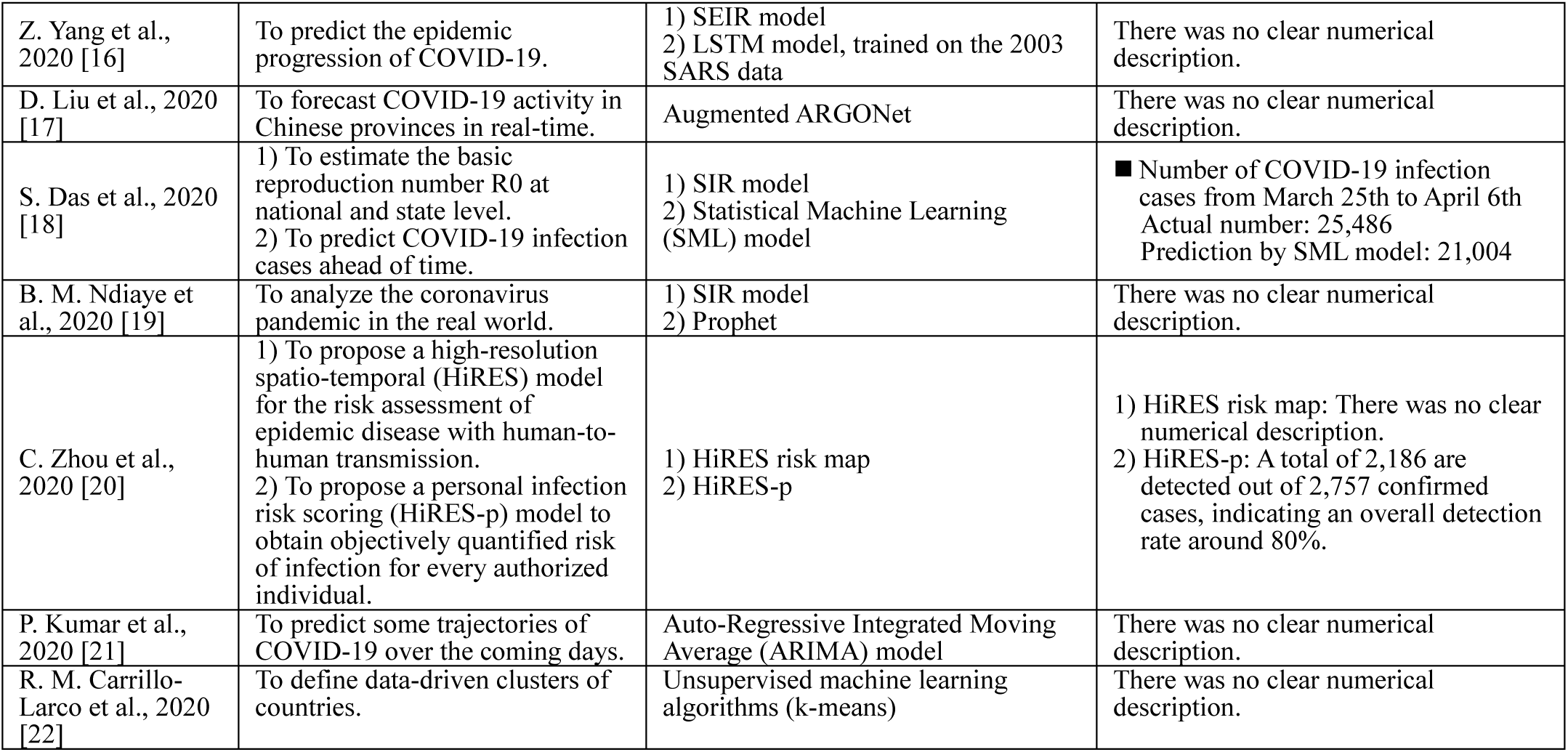
| Introduction of the existing works about ML models which predict the spreading of COVID-19 infection

By comparing the above existing works with our ML model, we found the following novelty points of our ML model.

- The below two additional items were input to our ML model as binary label (0 or 1). The details are described in **Supplementary Table 2**.

1. special measures taken by South Korean government to prevent the spreading of COVID-19 infection
2. date of South Korean legislative election
- By using the XGBoost in combination with the MultiOutputRegressor, multiple objective variables, i.e., the number of COVID-19 infection cases in each of 17 provinces can be output.

## 3. Methods

- Our machine learning (ML) model is a combination of XGBoost [2] and MultiOutputRegressor [3].
- XGBoost Official web sight of XGBoost: https://xgboost.readthedocs.io/en/latest/ XGBoost is a distributed gradient boosting library implements machine learning algorithms under the Gradient Boosting framework. The original XGBoost is a regressor that can only estimate (output) a single objective variable (explained variable). However, by combinating XGBoost and MultiOutputRegressor, we can estimate multiple objective variables (multiple explained variables).
- MultiOutoutRegressor Official web sight of MultiOutoutRegressor: https://scikit-learn.org/stable/modules/generated/sklearn.multioutput.MultiOutputRegressor.html MultiOutputRegressor is one of Scikit-Learn modules that can estimate multiple objective variables
- Task of our ML model: regression of daily infection cases in each of 17 provinces over the coming 24 days
- Loss function for training our ML model: Mean squared error (MSE) of numbers of daily infection cases in 17 provinces of South Korea over the coming 24 days
- Evaluation index for testing: binary classification (whether the ML model can classify provinces where total infection cases over the coming 24 days is more than 100).
- Hyper parameters:

I. max_depth: 6
II. n_estimators: 30
III. The rest of all hyper parameters: default (standard)

## 4. Dataset

- Dataset name: Data Science for COVID-19 in South Korea (DS4C)
- Contents: Information of 3,288 COVID-19 infection cases confirmed in South Korea from January 20th to April 29th, 2020. We decided to use this dataset since it contained more detailed information compared to other countries’ data. Note that we used only 3,285 cases apart from 3 cases with no mention of infection date.
- Dataset downloaded from: https://www.kaggle.com/kimjihoo/coronavirusdataset
- Dataset reported by: Korea Centers for Disease Control and Prevention (KCDC) and 17 provinces in South Korea
- License of the dataset: CC BY-NC-SA 4.0
- Training set: Information related to COVID-19 over the past 24 days (Jan. 25th - Feb. 14th) is input. Total increase in infection cases over the coming 24 days (Feb. 15th - Mar. 9th) in each of 17 provinces is output.
- Testing set: Information related to COVID-19 over the past 24 days (Feb. 15th - Mar. 9th) is input. Total increase in infection cases over the coming 24 days (Mar. 10th - Apr. 2th) in each of 17 provinces is output.
- COVID-19 information input to our ML model is shown in **Supplementary Tables 1 and 2**. In addition to the two tables, the number of daily infection cases in each province of South Korea over the past 24 days (e.g., the red box in **Fig. 1**) is input.

**Fig. 1.**
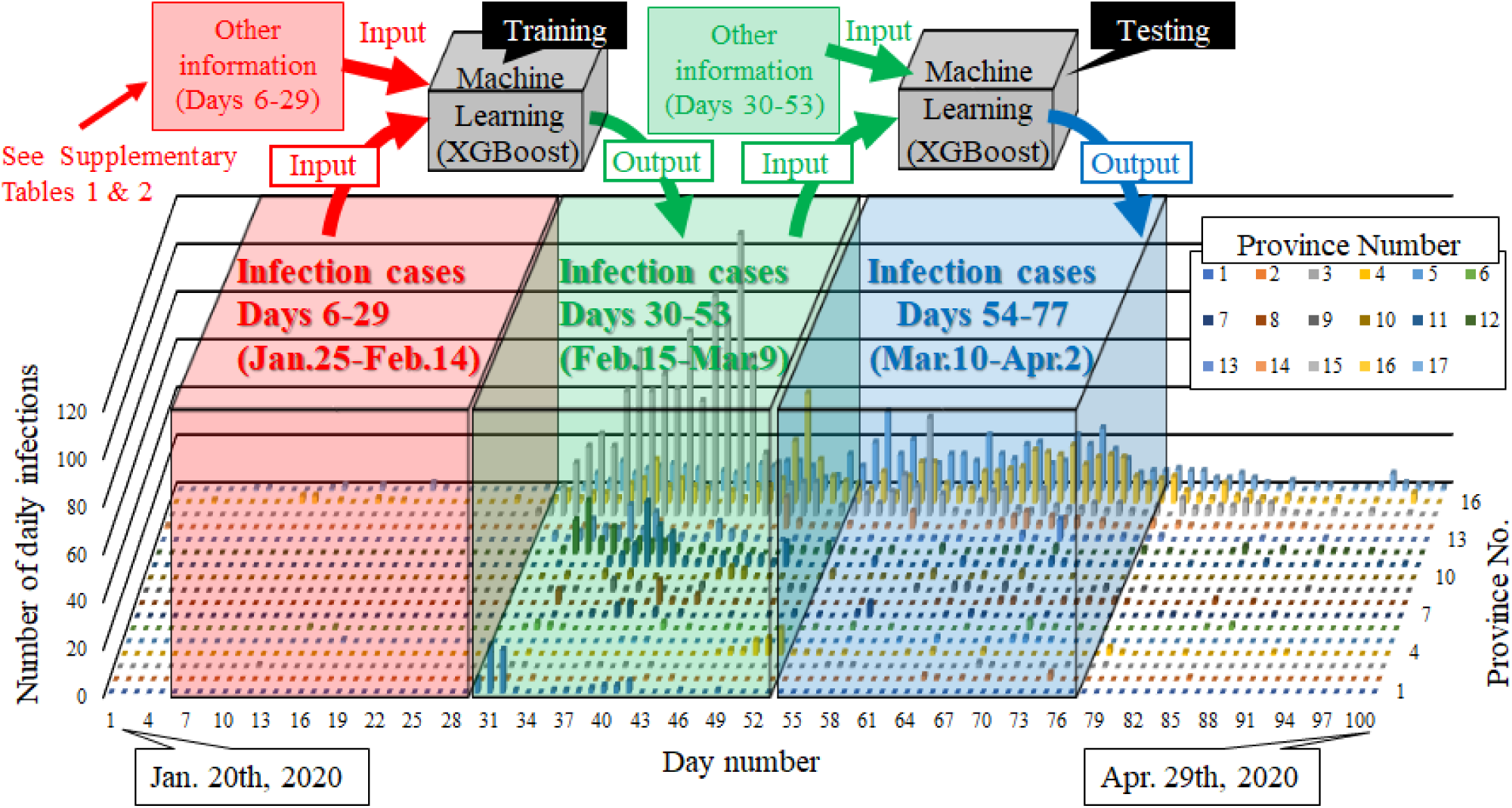
The COVID-19 dataset of South Korea used for training and testing our ML model (a combination of XGBoost and MultiOutputRegressor). This figure shows the number of daily infection cases in each of 17 provinces (South Korea, from January 20th to April 29th in 2020). January 20th is equal to Day 1 and April 29th is equal to Day 101. In training, the information related to COVID-19 over the past 24 days (Jan. 25th - Feb. 14th) is input. Then, the total increase in the number of infection cases over the coming 24 days (Feb. 15th - Mar. 9th) in each of 17 provinces is output. In testing, the information related to COVID-19 over the past 24 days (Feb. 15th - Mar. 9th) is input. Then, the total increase in the number of infection cases over the coming 24 days (Mar. 10th - Apr. 2th) in each of 17 provinces is estimated. The estimation performances of our ML model for the test set are shown in **Fig. 3**.

**Fig. 2.**
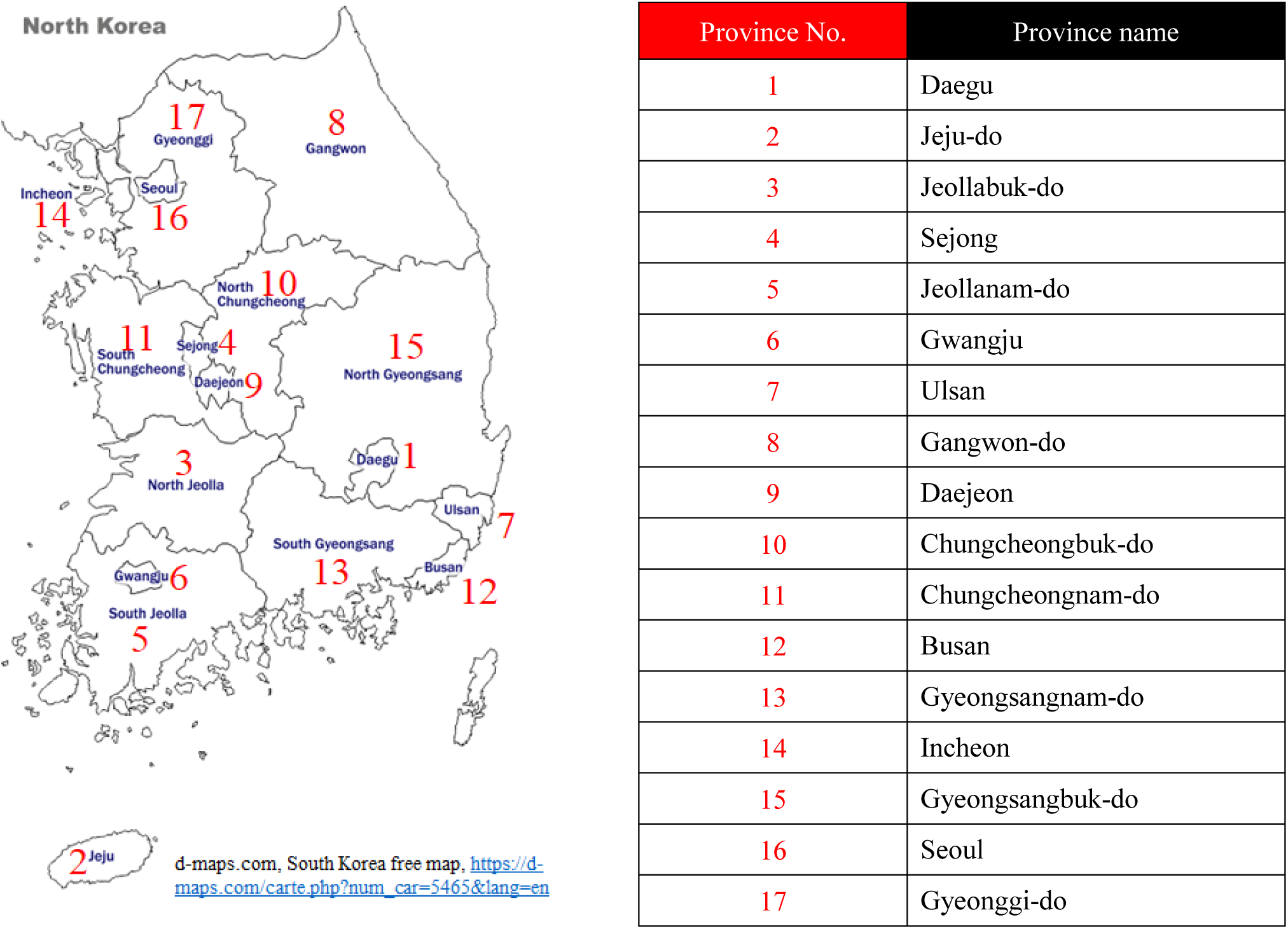
Province number allocated for each of 17 Provinces in South Korea.

**Fig. 3.**
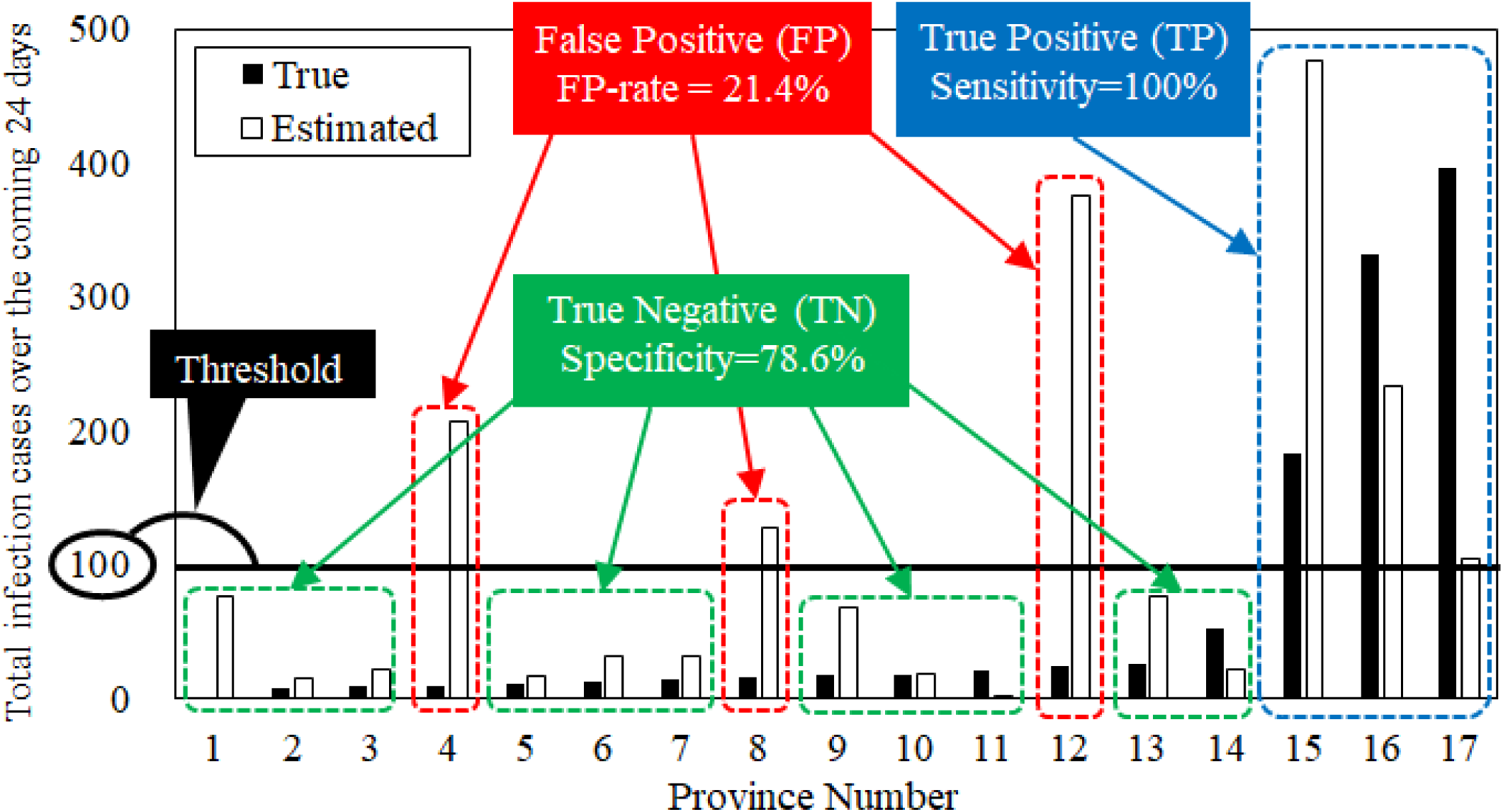
Performance validation (test) result of our ML model. The number of daily infection cases and the other information related to COVID-19 over the past 24 days (Feb. 15th - Mar. 9th) in each of 17 provinces is input. Then, the total increase in the number of infection cases over the coming 24 days (Mar. 10th - Apr. 2th) in each of 17 provinces is estimated. The black bar graph is the true value, and the white bar graph is the estimated value.

## 5. Results and discussion

The accuracy of the binary classification whether our ML model can classify provinces where total infection cases over the coming 24 days is more than 100 is as follows.

Sensitivity = TP / (TP + FN) = 3/3 = 100% (i.e., true positive rate, recall, hit rate)

Specificity = TN / (TN + FP) = 11/14 = 78.6% (i.e., true negative rate)

False Positive Rate = FP / (TN + FP) = 21.4%

Accuracy = (TP + TN) / (TP + TN + FP + FN) = 14/17 = 82.4 %

where,

TP = True Positive

TN = True Negative

FP = False Positive

FN = False Negative

Sensitivity=100% of the binary classification means that we did not overlook the three provinces where the number of COVID-19 infection cases increased by more than 100. In addition, as for the provinces where the actual number of new COVID-19 infection cases is less than 100, the ratio (Specificity) that our ML model can correctly estimate is 78.6%, which is relatively high.

Next, we evaluate the accuracy of our ML model from the perspective of the regression task, not the binary classification. The ratio that our ML model can estimate the increase of COVID-19 infection cases in each province over the coming 24 days when the maximum permissible error is set to 100 infection cases is as follows.

Another accuracy when the maximum permissible error is set to 100 infection cases = 12/17 = 70.6%

From the above all, it is demonstrated that there is a sufficient possibility that our ML model can support the following four points.

1. Promotion of behavior modification of residents in dangerous areas
2. Assistance for decision to resume economic activities in each region
3. Assistance in determining infectious disease control measures in each region
4. Search for factors that are highly correlated with the future increase in the number of COVID-19 infection cases.

### Notes for the performance of our ML model

- It is pointed out that the actual number of positives may be higher than this dataset because PCR tests may be insufficient. If this point is correct, there is a possibility that the performance of our ML model (sensitivity = 100%, specificity = 78.6%, false Positive Rate = 21.4%) may change.
- There is a possibility that the current input information may not contain important information. For example, in this method, population, population density, temperature, humidity, weather, regulation of economic activity, degree of land (urban area, depopulated area, industrial area, forest area), etc. of each province are not input. By inputting this information, the estimation performance might be improved.
- In this paper, both input and output of our ML model are set to 24 days, but this is not always optimal. Performance may change by changing the number of days.

## 6. Conclusions

We built a machine learning model (ML model) which input the number of daily infection cases and the other information related to COVID-19 over the past 24 days in each of 17 provinces in South Korea, and output the total increase in the number of infection cases in each of 17 provinces over the coming 24 days. We employ a combination of XGBoost and MultiOutputRegressor as machine learning model (ML model). We trained the ML model by setting loss function as the number of daily infections in the 17 provinces (i.e., regression task). We tested the ML model in terms of binary classification (whether the ML model can classify provinces where total infection cases over the coming 24 days is more than 100). As a result, Sensitivity = 3/3 = 100%, Specificity = 11/14 = 78.6%, False Positive Rate = 3/11 = 21.4%, Accuracy = 14/17 = 82.4 %. Sensitivity = 100% means that we did not overlook the three provinces where the number of COVID-19 infection cases increased by more than 100. In addition, as for the provinces where the actual number of new COVID-19 infection cases was less than 100, the ratio (Specificity) that our ML model could correctly estimate is 78.6%, which is relatively high. From the above all, it is demonstrated that there is a sufficient possibility that our ML model can support the following four points.

1. Promotion of behavior modification of residents in dangerous areas
2. Assistance for decision to resume economic activities in each region
3. Assistance in determining infectious disease control measures in each region
4. Search for factors that are highly correlated with the future increase in the number of COVID-19 infection cases.

## Data Availability

- Dataset name: Data Science for COVID-19 in South Korea (DS4C)
- Dataset downloaded from: https: //www.kaggle.com/kimjihoo/coronavirusdataset
- Dataset reported by: Korea Centers for Disease Control and Prevention (KCDC) and 17 provinces in South Korea
- License of the dataset: CC BY-NC-SA 4.0

## 9. Contributions of authors

Ayaka Suzuki, the corresponding author, contributed to the software engineering and developed all the codes.

Yoshiro Suzuki managed the project, analyzed the results, and wrote the paper.

The other authors helped designing this research project. Their contributions are almost equal to each other.

**Supplementary Table 1.**
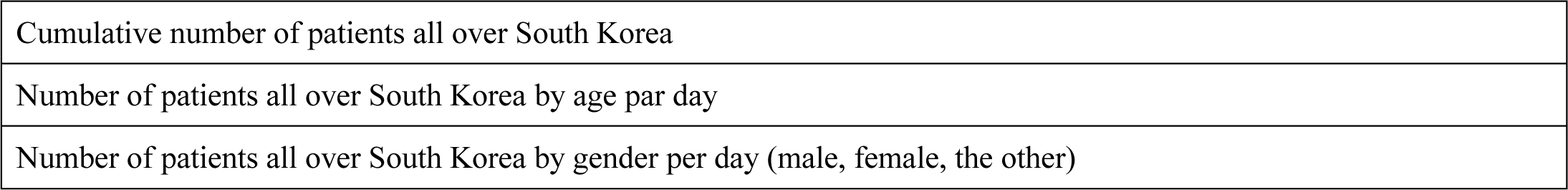
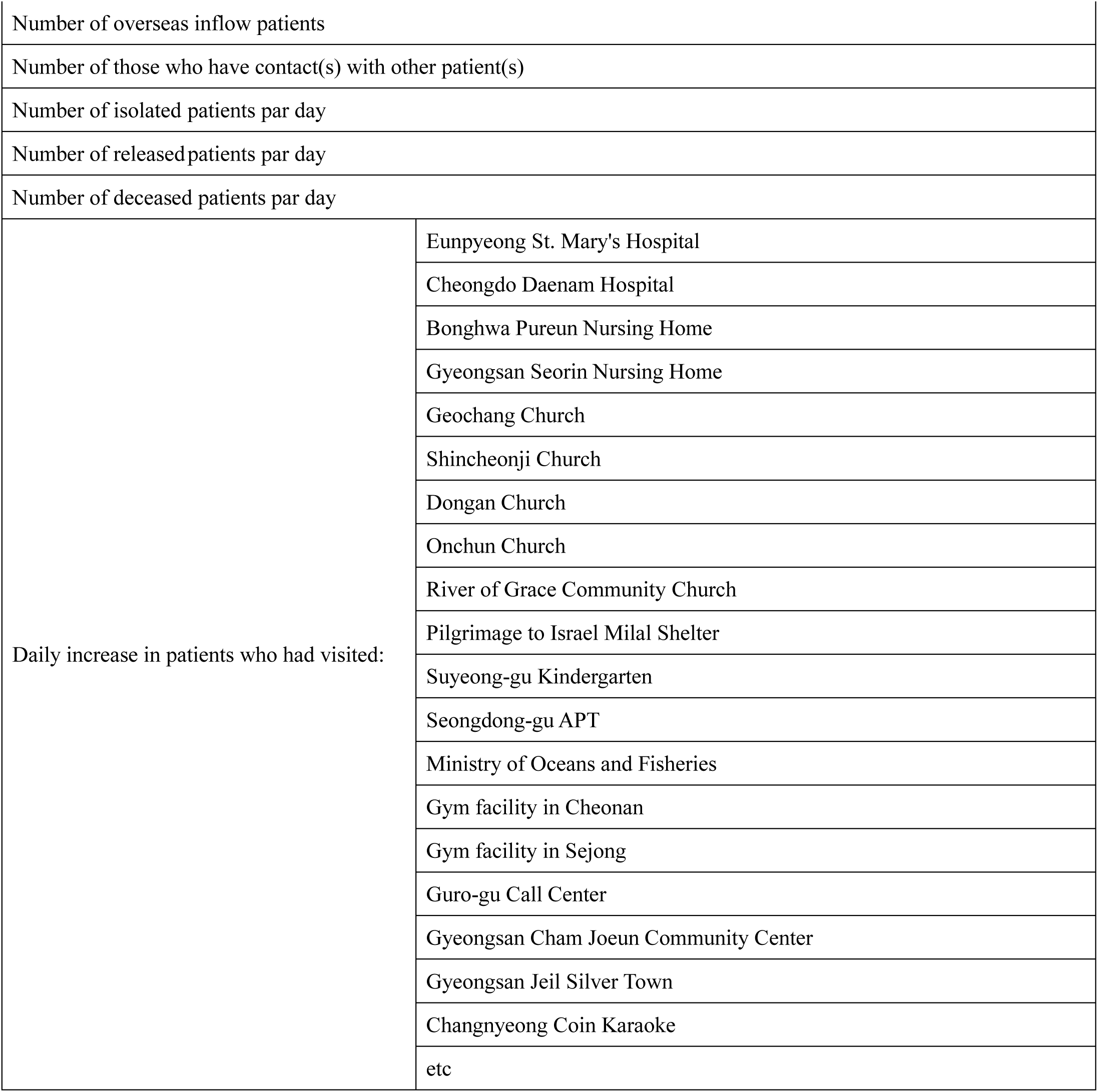
Natural number (NOT label) information input to our ML model. In addition to the information in this table, both information shown in **Supplementary Table 2** and information depicted in **Fig. 1** are input to our ML model.

|  |
| --- |
| Cumulative number of patients all over South Korea |
| Number of patients all over South Korea by age par day |
| Number of patients all over South Korea by gender per day (male, female, the other) |

|  |  |
| --- | --- |
| Number of overseas inflow patients |  |
| Number of those who have contact(s) with other patient(s) |  |
| Number of isolated patients par day |  |
| Number of released patients par day |  |
| Number of deceased patients par day |  |
| Daily increase in patients who had visited: | Eunpyeong St. Mary's Hospital |
|  | Cheongdo Daenam Hospital |
|  | Bonghwa Pureun Nursing Home |
|  | Gyeongsan Seorin Nursing Home |
|  | Geochang Church |
|  | Shincheonji Church |
|  | Dongan Church |
|  | Onchun Church |
|  | River of Grace Community Church |
|  | Pilgrimage to Israel Milal Shelter |
|  | Suyeong-gu Kindergarten |
|  | Seongdong-gu APT |
|  | Ministry of Oceans and Fisheries |
|  | Gym facility in Cheonan |
|  | Gym facility in Sejong |
|  | Guro-gu Call Center |
|  | Gyeongsan Cham Joeun Community Center |
|  | Gyeongsan Jeil Silver Town |
|  | Changnyeong Coin Karaoke |
|  | etc |

**Supplementary Table 2.**
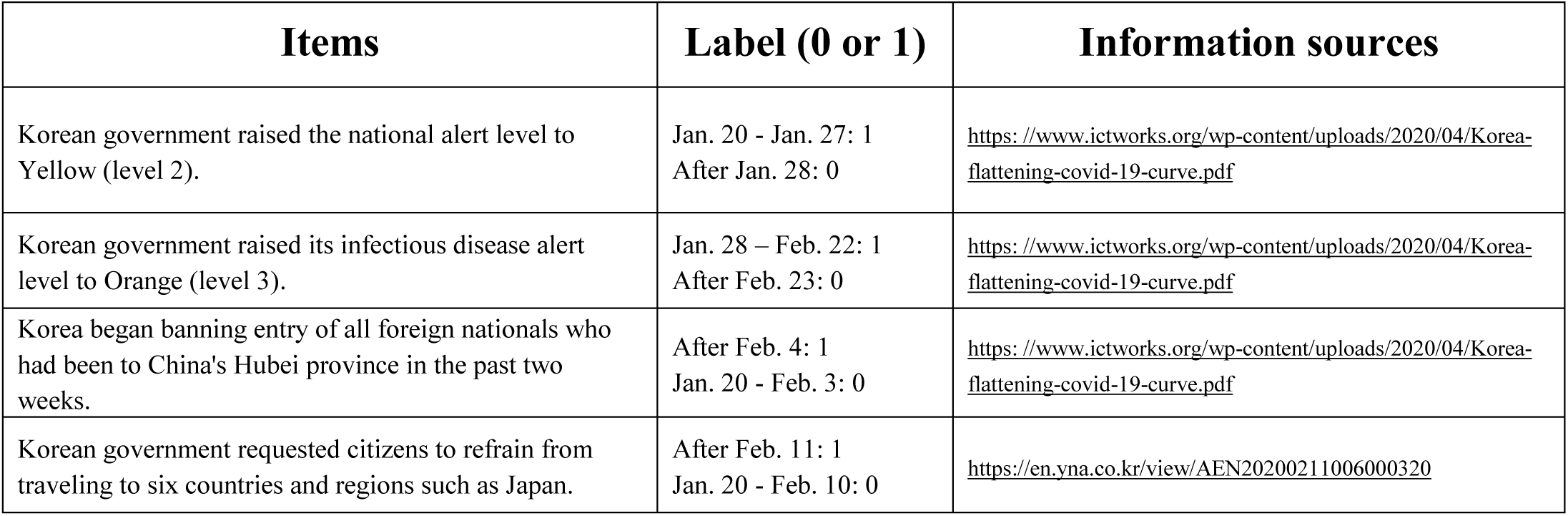
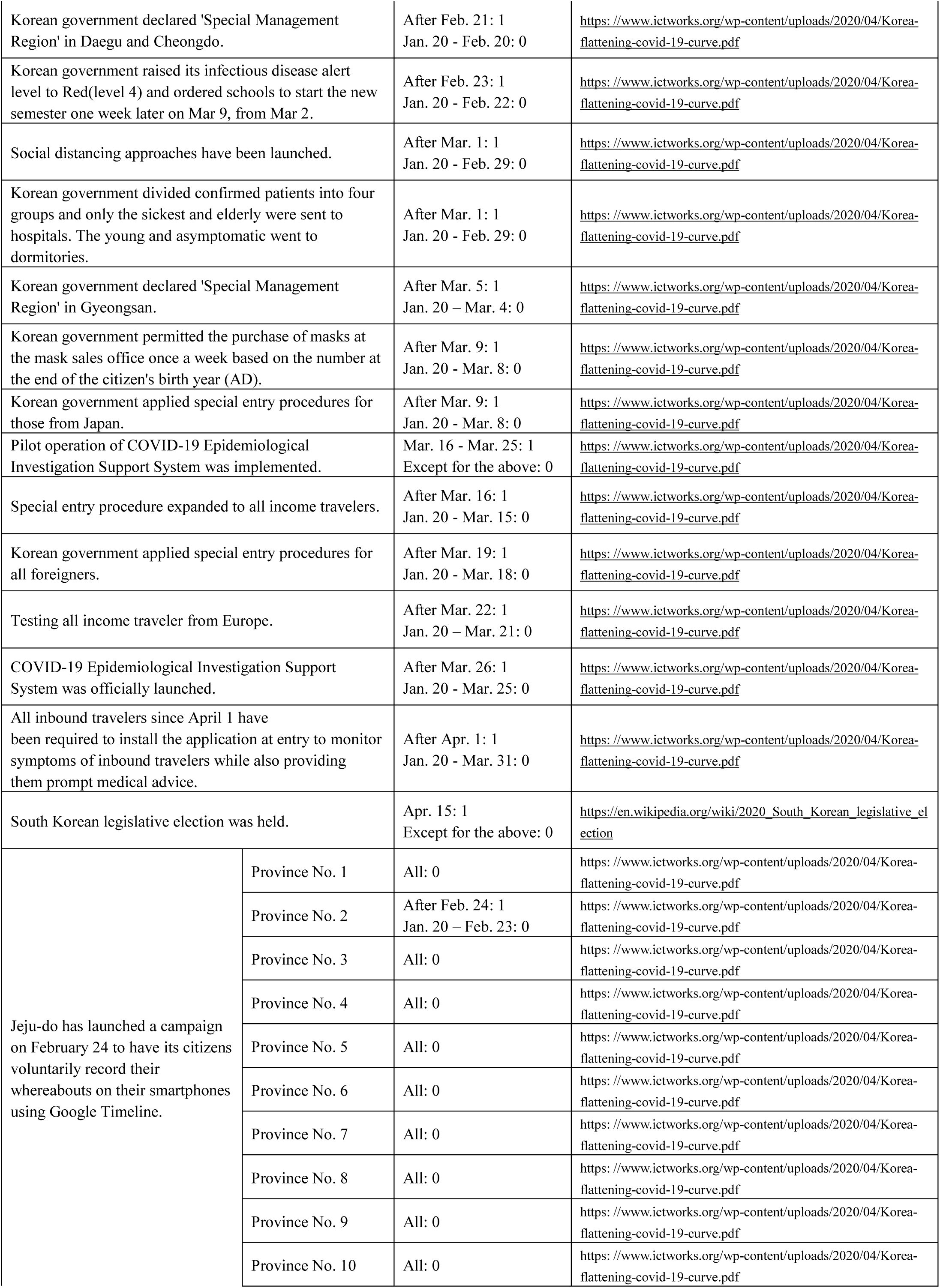
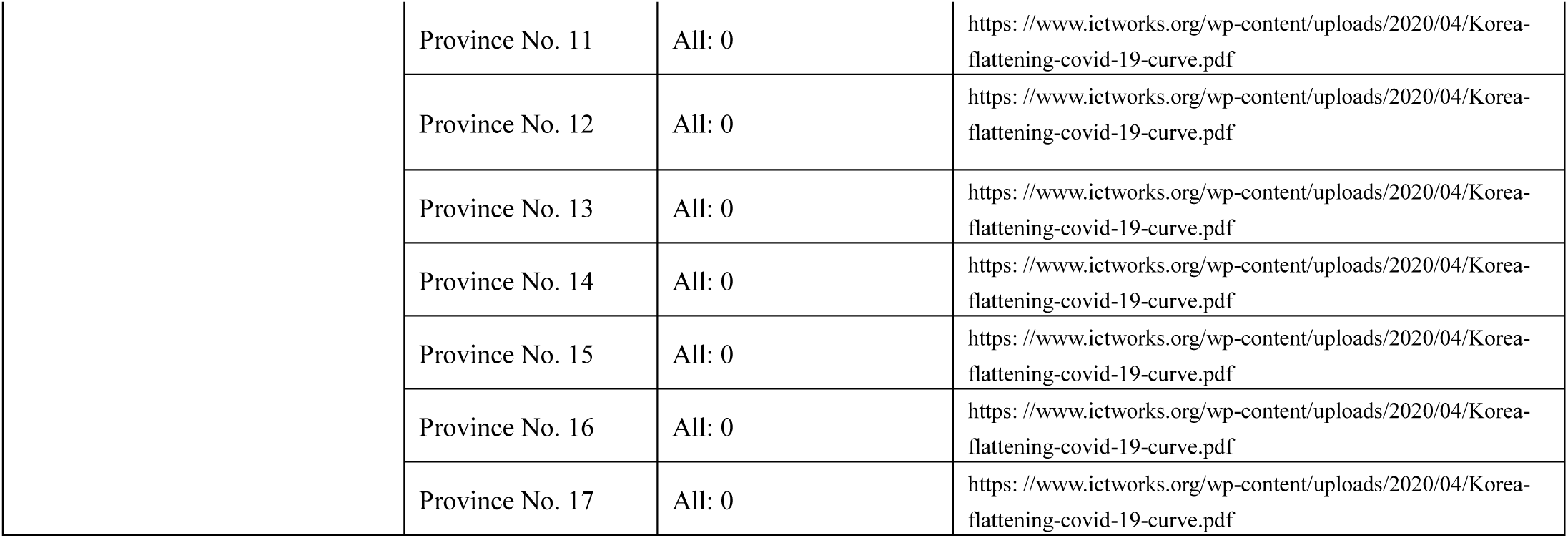
Label (NOT variable) information input to our ML model. In addition to the information in this table, both information shown in **Supplementary Table 1** and information depicted in **Fig. 1** are input to our ML model.

| Items | Label (0 or 1) | Information sources |
| --- | --- | --- |
| Korean government raised the national alert level to Yellow (level 2). | Jan. 20 - Jan. 27: 1<br>After Jan. 28: 0 | https://www.ictworks.org/wp-content/uploads/2020/04/Korea-flattening-covid-19-curve.pdf |
| Korean government raised its infectious disease alert level to Orange (level 3). | Jan. 28 – Feb. 22: 1<br>After Feb. 23: 0 | https://www.ictworks.org/wp-content/uploads/2020/04/Korea-flattening-covid-19-curve.pdf |
| Korea began banning entry of all foreign nationals who had been to China's Hubei province in the past two weeks. | After Feb. 4: 1<br>Jan. 20 - Feb. 3: 0 | https://www.ictworks.org/wp-content/uploads/2020/04/Korea-flattening-covid-19-curve.pdf |
| Korean government requested citizens to refrain from traveling to six countries and regions such as Japan. | After Feb. 11: 1<br>Jan. 20 - Feb. 10: 0 | https://en.yna.co.kr/view/AEN20200211006000320 |

|  |  |  |  |
| --- | --- | --- | --- |
| Korean government declared 'Special Management Region' in Daegu and Cheongdo. |  | After Feb. 21: 1<br>Jan. 20 - Feb. 20: 0 | https://www.ictworks.org/wp-content/uploads/2020/04/Korea-flattening-covid-19-curve.pdf |
| Korean government raised its infectious disease alert level to Red(level 4) and ordered schools to start the new semester one week later on Mar 9, from Mar 2. |  | After Feb. 23: 1<br>Jan. 20 - Feb. 22: 0 | https://www.ictworks.org/wp-content/uploads/2020/04/Korea-flattening-covid-19-curve.pdf |
| Social distancing approaches have been launched. |  | After Mar. 1: 1<br>Jan. 20 - Feb. 29: 0 | https://www.ictworks.org/wp-content/uploads/2020/04/Korea-flattening-covid-19-curve.pdf |
| Korean government divided confirmed patients into four groups and only the sickest and elderly were sent to hospitals. The young and asymptomatic went to dormitories. |  | After Mar. 1: 1<br>Jan. 20 - Feb. 29: 0 | https://www.ictworks.org/wp-content/uploads/2020/04/Korea-flattening-covid-19-curve.pdf |
| Korean government declared 'Special Management Region' in Gyeongsan. |  | After Mar. 5: 1<br>Jan. 20 - Mar. 4: 0 | https://www.ictworks.org/wp-content/uploads/2020/04/Korea-flattening-covid-19-curve.pdf |
| Korean government permitted the purchase of masks at the mask sales office once a week based on the number at the end of the citizen's birth year (AD). |  | After Mar. 9: 1<br>Jan. 20 - Mar. 8: 0 | https://www.ictworks.org/wp-content/uploads/2020/04/Korea-flattening-covid-19-curve.pdf |
| Korean government applied special entry procedures for those from Japan. |  | After Mar. 9: 1<br>Jan. 20 - Mar. 8: 0 | https://www.ictworks.org/wp-content/uploads/2020/04/Korea-flattening-covid-19-curve.pdf |
| Pilot operation of COVID-19 Epidemiological Investigation Support System was implemented. |  | Mar. 16 - Mar. 25: 1<br>Except for the above: 0 | https://www.ictworks.org/wp-content/uploads/2020/04/Korea-flattening-covid-19-curve.pdf |
| Special entry procedure expanded to all income travelers. |  | After Mar. 16: 1<br>Jan. 20 - Mar. 15: 0 | https://www.ictworks.org/wp-content/uploads/2020/04/Korea-flattening-covid-19-curve.pdf |
| Korean government applied special entry procedures for all foreigners. |  | After Mar. 19: 1<br>Jan. 20 - Mar. 18: 0 | https://www.ictworks.org/wp-content/uploads/2020/04/Korea-flattening-covid-19-curve.pdf |
| Testing all income traveler from Europe. |  | After Mar. 22: 1<br>Jan. 20 - Mar. 21: 0 | https://www.ictworks.org/wp-content/uploads/2020/04/Korea-flattening-covid-19-curve.pdf |
| COVID-19 Epidemiological Investigation Support System was officially launched. |  | After Mar. 26: 1<br>Jan. 20 - Mar. 25: 0 | https://www.ictworks.org/wp-content/uploads/2020/04/Korea-flattening-covid-19-curve.pdf |
| All inbound travelers since April 1 have been required to install the application at entry to monitor symptoms of inbound travelers while also providing them prompt medical advice. |  | After Apr. 1: 1<br>Jan. 20 - Mar. 31: 0 | https://www.ictworks.org/wp-content/uploads/2020/04/Korea-flattening-covid-19-curve.pdf |
| South Korean legislative election was held. |  | Apr. 15: 1<br>Except for the above: 0 | https://en.wikipedia.org/wiki/2020_South_Korean_legislative_election |
| Jeju-do has launched a campaign on February 24 to have its citizens voluntarily record their whereabouts on their smartphones using Google Timeline. | Province No. 1 | All: 0 | https://www.ictworks.org/wp-content/uploads/2020/04/Korea-flattening-covid-19-curve.pdf |
|  | Province No. 2 | After Feb. 24: 1<br>Jan. 20 - Feb. 23: 0 | https://www.ictworks.org/wp-content/uploads/2020/04/Korea-flattening-covid-19-curve.pdf |
|  | Province No. 3 | All: 0 | https://www.ictworks.org/wp-content/uploads/2020/04/Korea-flattening-covid-19-curve.pdf |
|  | Province No. 4 | All: 0 | https://www.ictworks.org/wp-content/uploads/2020/04/Korea-flattening-covid-19-curve.pdf |
|  | Province No. 5 | All: 0 | https://www.ictworks.org/wp-content/uploads/2020/04/Korea-flattening-covid-19-curve.pdf |
|  | Province No. 6 | All: 0 | https://www.ictworks.org/wp-content/uploads/2020/04/Korea-flattening-covid-19-curve.pdf |
|  | Province No. 7 | All: 0 | https://www.ictworks.org/wp-content/uploads/2020/04/Korea-flattening-covid-19-curve.pdf |
|  | Province No. 8 | All: 0 | https://www.ictworks.org/wp-content/uploads/2020/04/Korea-flattening-covid-19-curve.pdf |
|  | Province No. 9 | All: 0 | https://www.ictworks.org/wp-content/uploads/2020/04/Korea-flattening-covid-19-curve.pdf |
|  | Province No. 10 | All: 0 | https://www.ictworks.org/wp-content/uploads/2020/04/Korea-flattening-covid-19-curve.pdf |
| Province No. 11 | All: 0 | https://www.ictworks.org/wp-content/uploads/2020/04/Korea-flattening-covid-19-curve.pdf |
| Province No. 12 | All: 0 | https://www.ictworks.org/wp-content/uploads/2020/04/Korea-flattening-covid-19-curve.pdf |
| Province No. 13 | All: 0 | https://www.ictworks.org/wp-content/uploads/2020/04/Korea-flattening-covid-19-curve.pdf |
| Province No. 14 | All: 0 | https://www.ictworks.org/wp-content/uploads/2020/04/Korea-flattening-covid-19-curve.pdf |
| Province No. 15 | All: 0 | https://www.ictworks.org/wp-content/uploads/2020/04/Korea-flattening-covid-19-curve.pdf |
| Province No. 16 | All: 0 | https://www.ictworks.org/wp-content/uploads/2020/04/Korea-flattening-covid-19-curve.pdf |
| Province No. 17 | All: 0 | https://www.ictworks.org/wp-content/uploads/2020/04/Korea-flattening-covid-19-curve.pdf |

